# Prescription Cascades Associated with Acetylcholinesterase Inhibitors Use: A High-Throughput Sequence Symmetry Analysis

**DOI:** 10.1101/2025.11.24.25340868

**Authors:** Danielle Newby, Sai Sumedha Bobba, Berta Raventós, Elin Rowlands, Xihang Chen, Laura Molina Porcel, Carlen Reyes, Talita Duarte-Salles, Antonella Delmestri, Wai Yi Man, Edward Burn, Marti Catala Sabate, Nicole Pratt, Annika Jödicke, Daniel Prieto Alhambra

## Abstract

**Background:** Acetylcholinesterase inhibitors (AChEIs), prescribed for symptomatic dementia, are associated with adverse drug effects which may prompt new drug prescriptions, known as prescription cascades. We aimed to identify potential AChEIs-induced prescription cascades using high-throughput sequence symmetry analysis using real world data.

**Methods:** Patients aged 18+ with 365 days of prior observation initiating AChEIs (donepezil, rivastigmine and galantamine) were identified from Clinical Practice Research Datalink GOLD between 2002 to 2022. We screened 510 drug classes and 1,213 individual ingredients within ±180 days of AChEIs (365 days as sensitivity). Crude and adjusted sequence ratios (ASR) were calculated with 99% confidence intervals with positive signals undergoing review for clinical plausibility.

**Results:** We identified 66,155 initiators of AChEIs (median age 81 years [IQR 76 - 85]); 62.8% female). Of the ATC classes and individual ingredients evaluated, 51 and 46 signals were positive with 28 (55%) and 22 (48%) classified as potential prescription cascades after review, respectively. Prescriptions of new drugs acting on the gastrointestinal system showed positive signals such as antipropulsives (ASR 1.50 [1.28 - 1.75]), loperamide (ASR 1.52 [1.30 - 1.77]) and cyclizine (ASR 2.10 [1.72 - 2.59]).

Positive signals were found for drugs acting on the nervous system such as benzodiazepine derivatives (ASR 1.83 [1.55 - 2.16]) as well as those acting on the respiratory system such as corticosteroids (ASR 1.66 [1.33 - 2.08]), glucocorticoids (ASR 1.54 [1.30 - 1.83]) and ingredients such as beclomethasone (ASR 1.48 [1.21 - 1.82]). Most positive signals remained in sensitivity analysis, with additional signals mostly related to drugs acting on the nervous system.

**Conclusions:** Identified signals suggest potential prescription cascades related to AChEIs use corresponding to gastrointestinal, neuropsychiatric, dermatological and respiratory adverse drug events. While these findings require further validation, this study demonstrates the utility of high-throughput signal detection for identifying potential prescription cascades to support pharmacovigilance in high-risk populations.

**Key Points:**

- Prescription cascades occur where a new drug is prescribed to treat adverse effects caused by another drug; Acetylcholinesterase inhibitors (AChEI) may trigger such cascades due to their known adverse drug effects.
- This study identified potential prescription cascades following initiation of AChEI using high-throughput sequence symmetry analysis.
- Identified signals aligned with gastrointestinal, neuropsychiatric, dermatological, and respiratory adverse effects, supporting the biological plausibility of these prescription cascades.

**Why does this paper matter?:** This study provides real world evidence of potential prescription cascades associated with AChEI use, highlighting how adverse drug effects may drive prescribing and polypharmacy in dementia patients. By applying high-throughput signal detection, this work supports pharmacovigilance and safer prescribing practices for older adults.

## INTRODUCTION

Dementia is a growing public health challenge affecting over 57 million people worldwide, with the prevalence projected to grow to 153 million people by 2050(1).

Acetylcholinesterase inhibitors (AChEIs) (donepezil, rivastigmine, and galantamine) enhance neurotransmission by preventing the breakdown of acetylcholine within synapses. They temporarily stabilize cognitive function and delay progression of dementia related to neurodegenerative diseases such as Alzheimer’ disease and Parkinson’s disease(2). However, due to their increased stimulation of the parasympathetic nervous system, AChEIs are associated with a range of adverse drug events (ADEs), including gastrointestinal, urinary, neuropsychiatric, and cardiovascular disturbances(3,4). Due to their use as first-line treatments for dementia, millions of patients are potentially introduced to the risk of prescription cascades(5,6).

Prescription cascades can occur when an ADE from a prescribed drug is misinterpreted as a new medical condition, prompting the initiation of additional medications to manage the symptoms(7). This polypharmacy can lead to increased healthcare costs and higher risks of further ADEs, hospitalization and death(8,9), particularly in older adults vulnerable to drug-related harms such as those with dementia(10). Despite the significance of prescription cascades in clinical practice, there is limited research quantifying these events in the context of dementia treatments, specifically among users of AChEIs using real world data.

Sequence Symmetry Analysis (SSA) is a type of self-controlled, case-only screening analysis used for signal detection. It identifies potential associations between two drug events, typically an index drug and a marker drug by comparing the order of these events within individuals, making it useful for detecting prescription cascades and temporal relationships between treatments(11,12). SSA is particularly well-suited for detecting drug-related prescribing patterns in real-world data(13–15), allowing identification of known ADEs as well as rarer or unknown ADEs(16). However, to date, no studies have systematically applied this method to investigate prescription cascades involving AChEIs.

The aim of this study is to identify prescription cascades associated with the use of AChEIs using primary care data from the United Kingdom (UK). By employing SSA across both the drug class and ingredients level, we seek to enhance understanding of the downstream prescribing consequences of AChEIs and highlight opportunities for mitigating the risks of prescription cascades including polypharmacy and drug-drug interactions in this high-risk population.

## METHODS

### Study design, setting, and data sources

We carried out SSA, a case-only screening analysis, using routinely collected primary care data from the UK. People with a prescription for AChEIs [donepezil, galantamine, and rivastigmine] were identified from the CPRD GOLD database which includes data from GP practices representing ∼4.3 % of the UK population(17). CPRD GOLD established in 1987 contains pseudonymized patient-level information on demographics, lifestyle data, clinical diagnoses, prescriptions, and preventive care. To date it covers >21.3 million patients with nearly three million alive and currently registered in contributing GP practices. CPRD GOLD data was mapped to the Observational Medical Outcomes Partnership (OMOP) Common Data Model(18).

### Study participants

Patients were included if they were 18 years or older with a prescription for the index drugs (AChEIs). From these patients, only those with a specific marker of interest (Anatomical Therapeutic Chemical (ATC) class or ingredient) within the last 180 days before or after the index initiation between the study period (January 2003 to January 2023). Only those with 365 days of prior history based on the first prescription of the index and markers were included with a 365-washout applied. Patients were excluded if the date of prescription for the index and marker were on the same date.

### Marker definitions

Marker drugs were identified at the drug class level based on the ATC 4th level for the class level analysis (n = 909), and at the ingredient level for the drug level analysis (n = 31,556). We did not include ATC drug classes related to vaccines, vitamins, herbal medicines, food, beverages, supplements, tobacco, and lab tests. For individual drug ingredients, we included both pharmacologically active agents and non-pharmacological substances that may be prescribed (e.g., water, sodium chloride). All ingredients were retained as they may represent markers of clinical events, supportive care, or formulation components.

Drug classes and ingredients with less than 500 patient records were excluded. We also excluded the ATC classes and ingredients representing other anti-dementia drugs. This resulted in 510 drug classes and 1,213 drug ingredients initially available for analysis. A summary of numbers of excluded classes and drug ingredients can be found in the supplement (**Table S1-S2**).

### Statistical methods

We used the R package “CohortSymmetry”(19) to perform SSA to test each ATC class and ingredient in a high throughput manner. For each AChEIs (index)-marker pair, we determined the crude sequence ratio (CSR) as the number of patients who initiated the marker class/ingredient after AChEIs initiation divided by the number of patients who initiated the marker before the AChEIs. Excess initiation of a marker after the AChEIs, relative to before the AChEI, results in a CSR > 1 and may indicate the presence of a prescription cascade. We required the marker initiation to occur within 180 days prior or following AChEIs initiation for the primary analysis with a sensitivity analysis using a longer initiation window of ±365 days.

To account for changes in prescribing trends of index and marker over time, the null sequence ratio (NSR) was calculated for each index-marker pair. Briefly, NSR is the expected sequence ratio in the absence of any causal relationship between the marker and index drug, based on population-level prescribing trends(12,20). The adjusted sequence ratio (ASR) was calculated by dividing the CSR by the NSR for each pair to adjust for background prescribing trends. All ASRs with a lower 99% confidence interval > 1 were considered a positive prescription cascade signal.

To evaluate the performance of SSA before the main study, we first tested positive and negative control drug pairs: amiodarone followed by levothyroxine (positive control) and amiodarone followed by allopurinol (negative control). Additionally, we included a study specific positive control, AChEIs followed by memantine, to reflect a clinically expected treatment sequence in dementia care(21).

Baseline characteristics of patients including age, sex, specific AChEIs medication, and a predefined list of comorbidities and medications was characterized 180 to 1 day prior to AChEIs initiation(22,23).

All positive signals in the primary analysis were manually reviewed to differentiate potential prescription cascades from false positive signals. False positive signals could be due to various factors such as detection biases, changes in prescription trends, disease progression and reverse causation(14). Manual review of positive signals first involved assessment of the CSRs, ASRs, and NSRs of each marker as well as temporal prescribing patterns over the study period. Secondly, literature searches were used to assess evidence for potential prescription cascades and their underlying mechanisms to support signal classification.

All analytical code for this study is publicly available on GitHub (https://github.com/oxford-pharmacoepi/DementiaPSSA). Analyses were conducted using R version 4.2.3.

## RESULTS

We identified 66,155 initiators of AChEIs who were eligible for inclusion. The baseline characteristics of the cohort are summarized in **Table 1**. The majority of AChEIs initiators were female (62.76%) with a median age of 81 years [IQR 76 - 85] (highest proportion of initiators aged 80 to 89 (48.4%)). Donepezil was the most prescribed AChEI (73.5%), followed by rivastigmine (17.9%) and galantamine (15.7%).

**Table 1:**
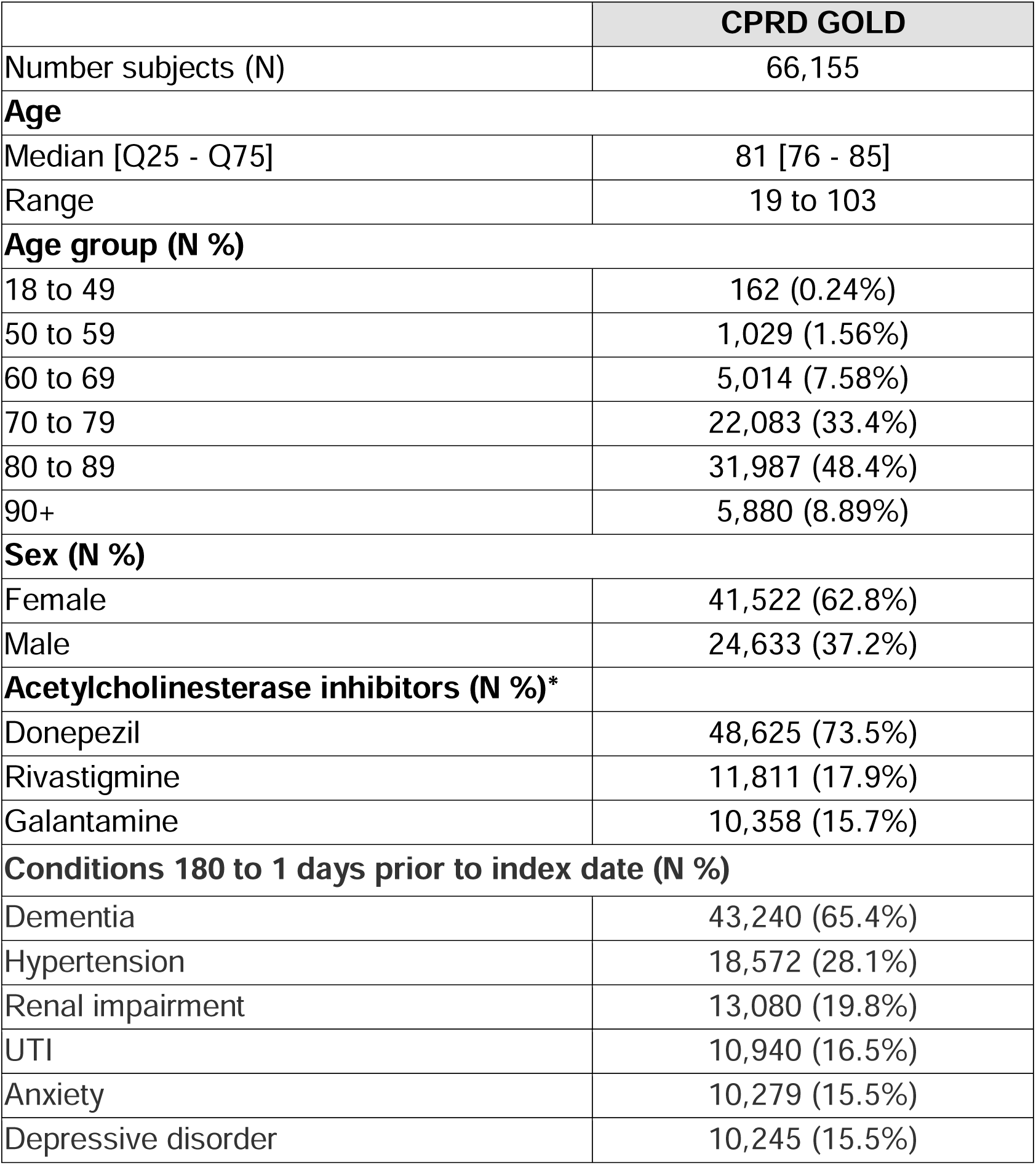
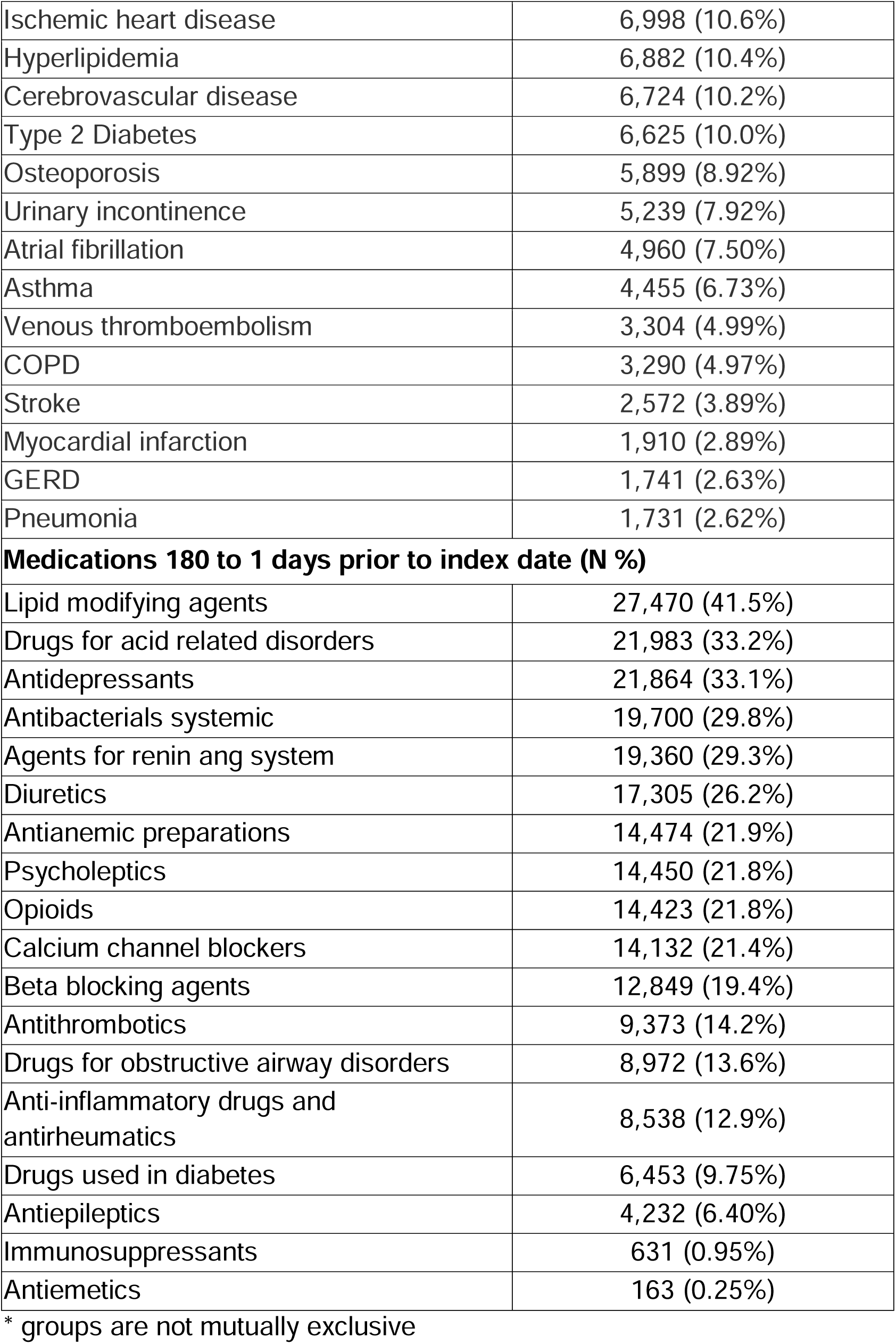
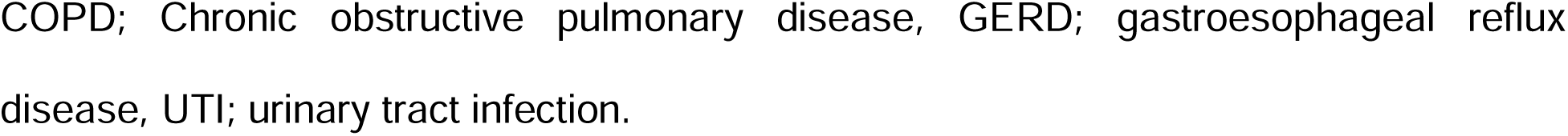
Baseline characteristics of acetylcholinesterase inhibitor initiators.

Within 180 days preceding AChEI initiation, 65.4% of patients had a dementia diagnosis. The most frequent comorbidities were hypertension (28.1%), renal impairment (19.8%), and urinary tract infections (16.5%). Commonly used medications included lipid-modifying agents (41.5%), drugs for acid-related disorders (33.2%), and antidepressants (33.1%).

All study results can be found in a user-friendly interactive web application (https://dpa-pde-oxford.shinyapps.io/dementia_pssa_study/).

The positive and negative controls were in line with expectations: the positive control amiodarone-levothyroxine drug pair showing a positive ASR of 5.33 [99% CI 4.52 - 6.33], while the negative control amiodarone-allopurinol pair showed a null signal (ASR 0.97 [0.76 - 1.24]). The study-specific positive control AChEIs-memantine pair also showed a positive ASR of 7.18 [5.92 - 8.79].

Among marker ATC drug classes analyzed, there were 51 significant signals identified in the 180-day primary analyses distributed across multiple ATC categories (**Figure 1**). Notable signals within the ATC alimentary tract and metabolism category included A04AD (other antiemetics) with an ASR of 3.64 (99% CI: 2.43 - 5.59), and A03FA (propulsives) with an ASR of 1.58 (1.33–1.87). In the nervous system ATC category, N07BC (drugs used in opioid dependence) had an ASR of 9.86 (5.06 - 21.6) where as N05CM (other hypnotics and sedatives) had an ASR of 2.77 (1.97 - 3.95). Additional signals were observed for N02AB (oripavine derivatives 1.35 [1.08 - 1.70]) and N02AB (phenylpiperidine derivatives 2.67 [1.82–3.99]). Several subgroups in the dermatological ATC class, notably corticosteroids such as D07AB (corticosteroids moderately potent group ii) and D07AC (corticosteroids potent group iii), showed ASRs of 1.20 (1.05 - 1.37) and 1.16 (1.02 - 1.33) respectively. Other signals were observed for systemic anti-infectives such as J01AA (tetracyclines) and J01CA (penicillins with extended spectrum) displayed positive signals. ATC classes from the respiratory system also showed positive signals including R01AD (corticosteroids for nasal use) and R03BA (glucocorticoids for obstructive airway diseases) had significant signals with ASRs of 1.66 (1.33 - 2.08) and 1.54 (1.30 - 1.83).

**Figure 1:**
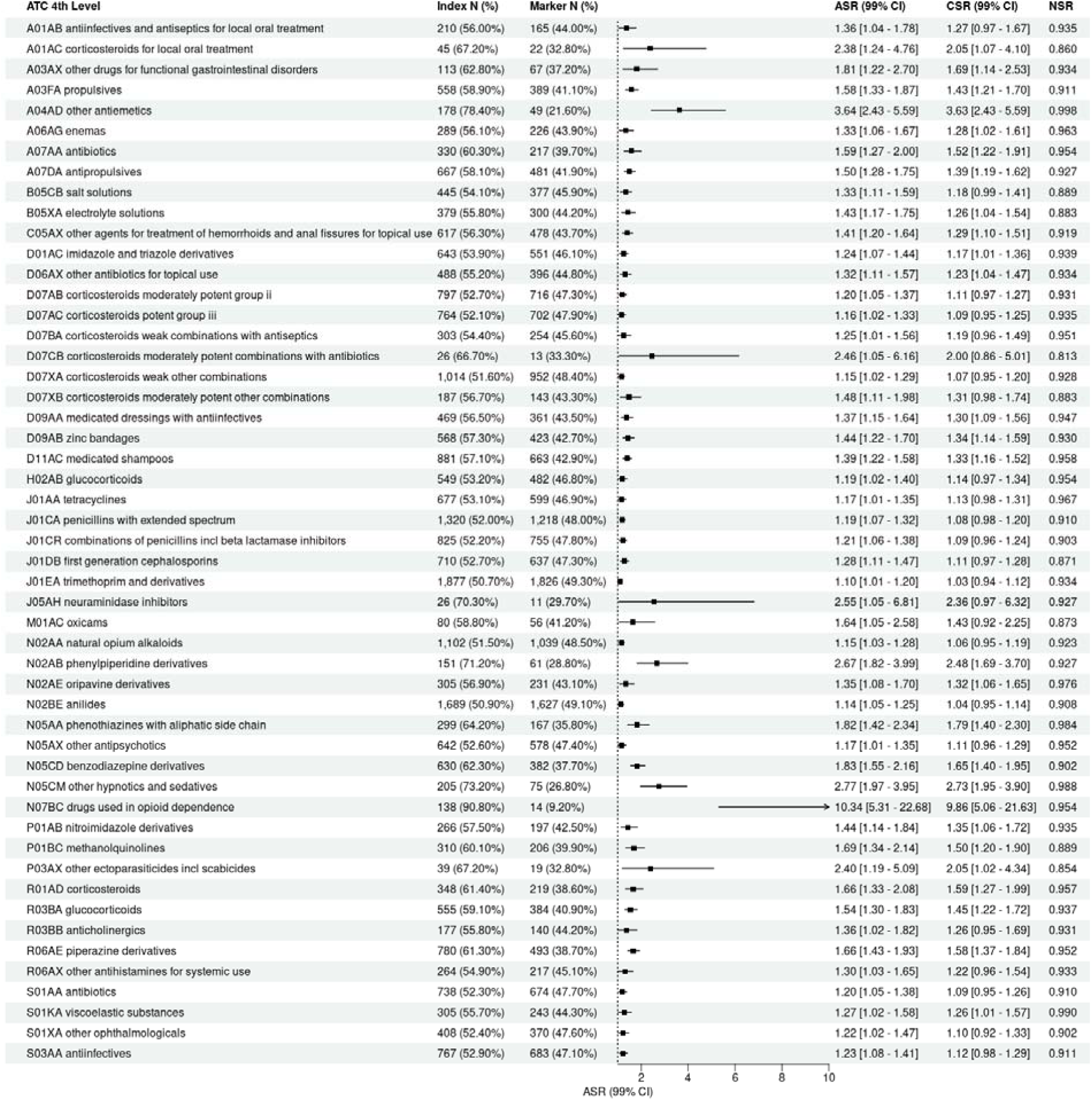
Forest Plot of positive signals from high-throughput sequence symmetry analysis of Acetylcholinesterase inhibitor versus ATC Drug Classes (4^th^ level) with a 180-day initiation window (ASR: adjusted sequence ratio, CSR: crude sequence ratio, NSR: null sequence ratio)

Negative signals (ASR <1) were also observed for some ATC classes (n = 16) including drugs for diabetes (A10BA, biguanides; A10BB, sulfonylureas), cardiovascular drugs (thiazides, ACE inhibitors, dihydropyridine derivatives, HMG-CoA reductase inhibitors), and several other classes such as antithrombotic agents, antianemic preparations, thyroid therapies, salicylic acid derivatives, certain anti-psychotics (diazepines/oxazepines/thiazepines/oxepines), and antidepressants (e.g. selective serotonin reuptake inhibitors) (**Table S3**).

Of the 51 positive signals, 42 (82%) remained positive when the index marker initiation window increased to 365 days, with an additional 17 positive signals not in the primary analysis (**Figure S1**). These additional positive signals included ATC classes for antiemetics (A04AA 1.67 [1.01 - 2.81]) and drugs for constipation (e.g. A06AA 1.15 [1.04 - 1.27]). Several nervous system drugs including antiepileptics (N03AG 1.52 [1.14 - 2.01]), anticholinergic agents (N04AA 1.65 [1.07 - 2.57]), antipsychotics (N05AD 1.54 [1.29 - 1.84], N05AL 1.31 [1.06 - 1.63]), anxiolytics (N05BA) and sedatives (N05CF, N05CH) were also seen as well as iron preparations (B03AB, B03AE), antibacterials (J01DC, J01FA) and nasal preparations (R01AX) (**Table S4**).

Among marker ingredients analyzed, there were 46 positive signals identified in the 180-day primary analyses (**Figure 2**). Positive signals spanned various ingredients including analgesics, gastrointestinal agents, sedatives, corticosteroids, and antipsychotics. Positive signals included opioids such as morphine (ASR 2.54 [2.03 - 3.20]), fentanyl (2.75 [1.87 - 4.13]), and buprenorphine (1.35 [1.08 - 1.70]). Hypnotics and sedatives such as midazolam (11.20 [6.89 - 19.39]) and methotrimeprazine (18.18 [8.25 - 48.64]) also were positive. Other ingredients included corticosteroids (beclomethasone 1.48 [1.21 - 1.82], fluticasone 1.51 [1.14 - 1.99]), antibiotics (trimethoprim 1.10 [1.01- 1.20], metronidazole 1.42 [1.15 -1.76], amoxicillin 1.20 [1.08 - 1.33]), and gastrointestinal agents such as domperidone (1.54 [1.24 - 1.91]) and loperamide (1.52 [1.30 - 1.77]). Non-pharmaceutical agents such as water (8.47 [5.07 - 15.11]) and sodium chloride (1.27 [1.08 - 1.51]) also showed positive signals.

**Figure 2:**
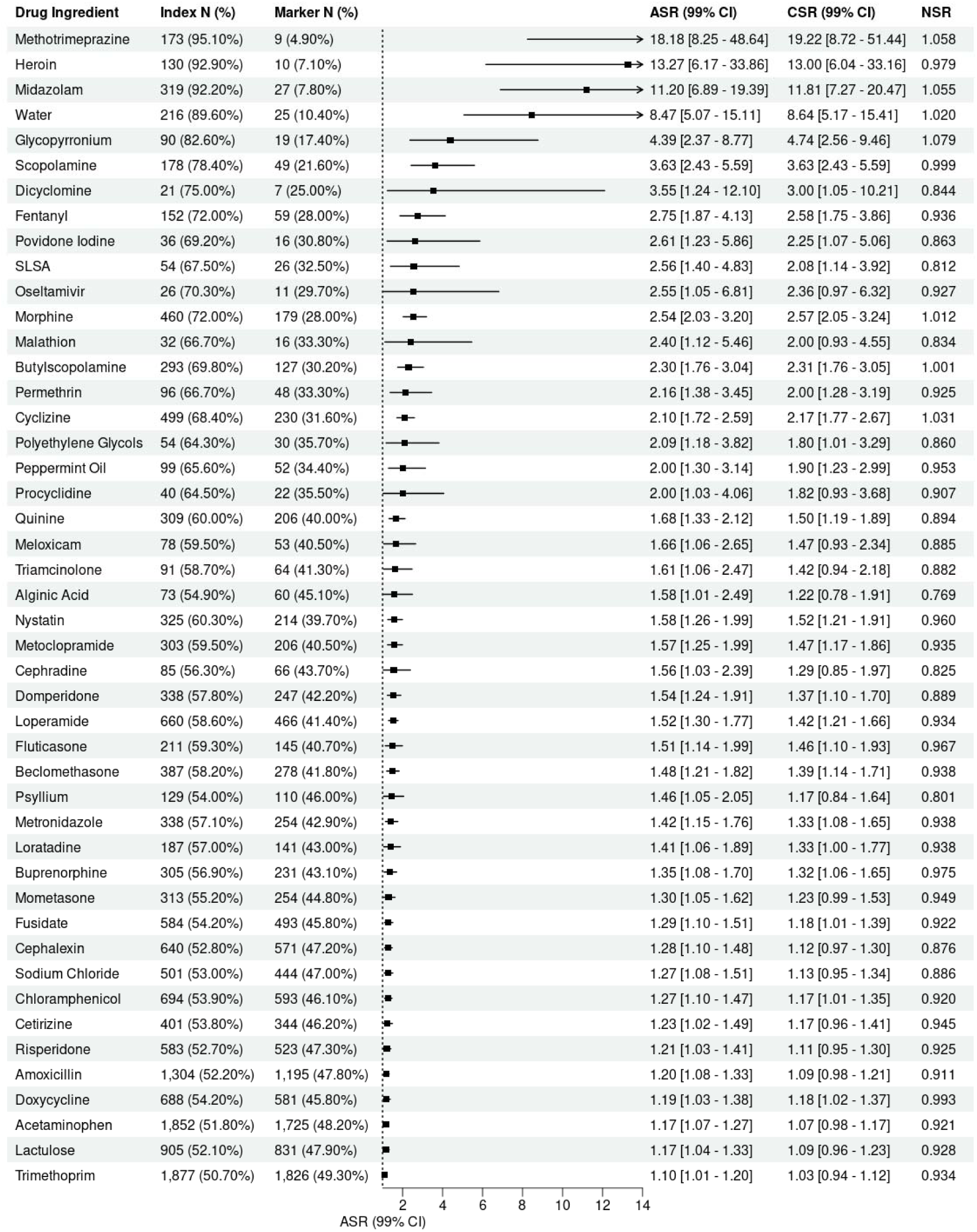
Forest Plot of positive signals from high-throughput sequence symmetry analysis of Acetylcholinesterase inhibitor versus drug ingredients with a 180-day initiation window (ASR: adjusted sequence ratio, CSR: crude sequence ratio, NSR: null sequence ratio)

Negative signals (n = 31) were again observed across several therapeutic classes including cardiovascular and anticoagulant agents (e.g., aspirin, amlodipine, apixaban), lipid-lowering therapies (e.g., atorvastatin, simvastatin), antidiabetic drugs (e.g., metformin, gliclazide), thyroid and hematological ingredients (e.g., levothyroxine, vitamin B12), and psychotropic medications (e.g., sertraline, quetiapine) (**Table S5**).

Increasing the initiation window to 365 days, 38 (83%) remained positive with 19 additional positive signals compared to the primary analysis (**Figure S2**). These additional positive signals included psychotropic and neurological drugs such as amisulpride (ASR 1.30 [1.04–1.64]), trazodone (1.44 [1.23–1.69]), valproate (1.51 [1.14–2.01]), haloperidol (1.53 [1.28–1.83]), and lorazepam (1.58 [1.41–1.77]). Other ingredients included antiemetics (ondansetron), laxatives (sennoside B, docusate), respiratory agents (ipratropium), and analgesics (oxycodone) (**Table S6**).

Review of positive signals across ATC classes and ingredients determined 28 (55%) and 22 (48%) signals were potential prescription cascades. The review of the positive signals and their prioritization can be found in the supplement (**Table S7 and S8**). The review of the additional positive signals from the 365-window sensitivity analysis is also provided in the supplement (**Table S9 and S10**).

## DISCUSSION

This high-throughput SSA identified several drug classes and ingredients whose initiation following AChEIs were consistent with possible prescription cascades, where side effects or complications from AChEIs prompted additional medication use. Many positive signals were linked to known adverse effects of AChEIs, particularly gastrointestinal, dermatological, neuropsychiatric, and respiratory systems.

AChEIs inhibit acetylcholinesterase, preventing the rapid breakdown of acetylcholine in the peripheral nervous system(24). The accumulation of acetylcholine leads to overstimulation of cholinergic receptions leading to several adverse events including diarrhea, nausea, vomiting, dizziness, muscle cramping, tremors, insomnia, urinary incontinence and seizures with evidence showing commonly reported ADEs are neuropsychiatric, gastrointestinal, and cardiovascular in nature(25). This pharmacological profile provides a plausible biological rationale for several positive signals observed in this work.

Increasing acetylcholine levels cause stimulation of muscarinic receptors in the nasal passages and airways, leading to increased mucus production, rhinorrhea, bronchial secretions, and bronchoconstriction(26,27); supported by this work and others with signals for medications used to counter these effects such as antihistamines, glucocorticoids and corticosteroids(28). Gastrointestinal related drugs reflect treatments for common cholinergic side effects such as nausea, vomiting, and motility disturbances(29,30). Furthermore, signals related to dermatological drugs and ingredients, including topical corticosteroids suggest management of drug-induced skin reactions(29), however, could be due to consequences of aging and associated care(31). Psychiatric conditions and treatments are frequently associated with dementia(32). Positive signals may reflect both side effects of AChEIs(33) and progression of the underlying disease(34). This interpretation is supported by our sensitivity analysis, which identified additional neurological medications using a wider initiation window. Consistent signals across both windows strengthen confidence in these associations, whereas discrepancies may also indicate differences between acute and delayed effects or prescribing practices related to disease progression.

Although signals obtained in this work align with known side effects some positive signals, such as systemic antibiotics and antiparasitic agents, are more likely attributable to comorbid infections rather than direct prescription cascades related to AChEIs initiation. However, there is conflicting evidence regarding AChEIs and infection risk(35,36). Furthermore, evidence shows AChEIs are associated with urinary incontinence leading to prescription of antimuscarinic medications, with no positive signals observed in our study. This could reflect under-ascertainment of events, variability in prescribing, awareness of polypharmacy and anticholinergic as well as individual AChEIs drug effects. A comparative effectiveness study showed higher prevalence of antimuscarinic prescriptions comparing donepezil to rivastigmine(37) whereas another study showed no difference between patients with dementia prescribed AChEIs with those not prescribed them(38). Similarly, a study identified an positive association between beta blockers and AChEI using prescription data from the Netherlands (39), whereas our analysis found a negative signal. This difference is likely due to the shorter initiation window of 180 days used for this work compared to the two years used by the other study. Longer initiation windows can give rise to time varying confounding with careful consideration of appropriate windows considering the disease specific context with sensitivity analysis required. Differences in population characteristics or prescribing patterns of AChEIs may also explain this discrepancy.

A small number of negative signals were observed in this study. While these should be interpreted cautiously, such findings could suggest either protective prescribing patterns, competing risks, or clinical avoidance of certain medications in patients receiving AChEIs, with clinical guidance recommending reviewing and deprescribing medications to prevent inappropriate prescription cascades in older populations(40).

Beyond acting as proxies for adverse effects, prescription cascades are also clinically important. The initiation of additional treatments to manage side effects contributes to polypharmacy, increasing the risk of drug–drug interactions, adverse drug events, and treatment burden(41). Drugs such as benzodiazepines, which may increase sedation and fall risk(42), gastrointestinal agents such as domperidone, which are associated with QT prolongation(43) showed positive signals and illustrate how prescription cascades may contribute to polypharmacy and drug-drug interactions following AChEI initiation. In older adults with dementia, such prescription cascades may further increase the risk of adverse events and contribute to cognitive decline(44).

Strengths of this study include the use of a large, comprehensive electronic health records database that enabled real-world pharmacoepidemiologic signal detection across a large population. The use of SSA reduced bias from time-invariant confounders and accounted for certain forms of time-varying confounding via adjustment of prescribing trends. Analyses were conducted at both the drug class (ATC level) and individual ingredient level, allowing identification of patterns that may be obscured at the class level and decreased power at the ingredient level offering complementary insights.

However, our study has some limitations. SSA is a signal detection method used to detect temporal asymmetry; therefore, it cannot be used to establish causality, with further validation of positive signals required using more robust epidemiological methods(45). Some signals may reflect disease progression rather than prescription cascades. For example, increased use of antipsychotics or sedatives could be driven by worsening dementia symptoms, which may occur following initiation of AChEIs(46,47). Prescription records in primary care identify if a medication was prescribed, but they do not confirm whether the patient took the drug or adhered to the treatment regimen. Additionally, as AChEIs are often initiated in specialist care before appearing in primary-care records, early adverse events or prescribing cascades that may occur might be missed leading to misclassification and attenuation of signals. We did not restrict our population to individuals over 65 years of age, based on the assumption that the biological processes underlying the development of side effects from AChEIs would not differ by age with most of the population in this study being older any attenuation due to including younger individuals was expected to be minimal. Finally, some positive signals may reflect confounding by indication, reverse causation and observational period imbalance.

## CONCLUSION

In summary, this study identified potential prescription cascades associated with initiation of AChEIs. Many associations corresponded with known ADEs, supporting the biological plausibility of these findings, while others may reflect disease progression or indirect prescribing behaviors. These findings demonstrate the value of high-throughput signal detection for identifying potential prescription cascades to help inform pharmacovigilance in high-risk populations. Further studies using complementary epidemiologic designs are warranted to confirm and validate findings.

## Data Availability

Patient level data used in this study was obtained through an approved application-the CPRD (application number 23_003334) and is only available following an approval process-safeguard the confidentiality of patient data. Details on how to apply for data access can be found at https://cprd.com/data-access.

## ACKNOWLEDGEMENTS

We thank Dr Ty Stanford for his initial statistical input and review of the manuscript.

## Author Contributions

Conceptualization; DN, DPA, EB; Data harmonisation and data quality assessment; AD, WYM; Formal analysis; DN; Funding acquisition; DPA; Supervision; DN; Interpretation of results: All authors; Roles/Writing - original draft: DN, SSB; Writing - review & editing: All authors

## DATA AVAILIABILITY STATEMENT

This study is based in part on data from the Clinical Practice Research Datalink (CPRD) obtained under licence from the UK Medicines and Healthcare products Regulatory Agency. The data is provided by patients and collected by the NHS as part of their care and support. The interpretation and conclusions contained in this study are those of the author/s alone. Patient level data used in this study was obtained through an approved application-the CPRD (application number 23_003334) and is only available following an approval process-safeguard the confidentiality of patient data. Details on how to apply for data access can be found at https://cprd.com/data-access.

Individual patient consent was not required, as CPRD data are de-identified and provided under ethical approval from the UK Health Research Authority (HRA) and the NHS Health and Social Care Research Ethics Committee.

## REFERENCES

1. Chen S, Cao Z, Nandi A, Counts N, Jiao L, Prettner K, et al. The global macroeconomic burden of Alzheimer’s disease and other dementias: estimates and projections for 152 countries or territories. Lancet Glob Health [Internet]. 2024 Sep 1 [cited 2025 Oct 14];12(9):e1534–43. Available from: https://www.thelancet.com/action/showFullText?pii=S2214109X2400264X

2. Marucci G, Buccioni M, Ben DD, Lambertucci C, Volpini R, Amenta F. Efficacy of acetylcholinesterase inhibitors in Alzheimer’s disease. Neuropharmacology [Internet]. 2021 Jun 1 [cited 2025 Oct 14];190:108352. Available from: https://www.sciencedirect.com/science/article/pii/S0028390820304202

3. Ruangritchankul S, Chantharit P, Srisuma S, Gray LC. Adverse Drug Reactions of Acetylcholinesterase Inhibitors in Older People Living with Dementia: A Comprehensive Literature Review. Ther Clin Risk Manag [Internet]. 2021 [cited 2025 Oct 14];17:927. Available from: https://pmc.ncbi.nlm.nih.gov/articles/PMC8427072/

4. Donepezil hydrochloride | Drugs | BNF | NICE [Internet]. [cited 2025 Oct 15]. Available from: https://bnf.nice.org.uk/drugs/donepezil-hydrochloride/

5. Bloomstone S, Anzuoni K, Cocoros N, Gurwitz JH, Haynes K, Nair VP, et al. Prescribing cascades in persons with Alzheimer’s disease: engaging patients, caregivers, and providers in a qualitative evaluation of print educational materials. Ther Adv Drug Saf [Internet]. 2020 [cited 2025 Oct 15];11:2042098620968310. Available from: https://pmc.ncbi.nlm.nih.gov/articles/PMC7675869/

6. Gromek KR, Thorpe CT, Aspinall SL, Hanson LC, Niznik JD. Anticholinergic co-prescribing in nursing home residents using cholinesterase inhibitors: potential deprescribing cascade. J Am Geriatr Soc [Internet]. 2022 Jan 1 [cited 2025 Oct 15];71(1):77. Available from: https://pmc.ncbi.nlm.nih.gov/articles/PMC9870851/

7. Dreischulte T, Shahid F, Muth C, Schmiedl S, Haefeli WE. Prescribing Cascades: How to Detect Them, Prevent Them, and Use Them Appropriately. Dtsch Arztebl Int [Internet]. 2022 Nov 22 [cited 2025 Oct 14];119(44):745. Available from: https://pmc.ncbi.nlm.nih.gov/articles/PMC9853235/

8. Leelakanok N, Holcombe AL, Lund BC, Gu X, Schweizer ML. Association between polypharmacy and death: A systematic review and meta-analysis. Journal of the American Pharmacists Association [Internet]. 2017 Nov 1 [cited 2025 Oct 14];57(6):729–738.e10. Available from: https://pubmed.ncbi.nlm.nih.gov/28784299/

9. Maher RL, Hanlon J, Hajjar ER. Clinical Consequences of Polypharmacy in Elderly. Expert Opin Drug Saf [Internet]. 2013 Jan [cited 2025 Oct 14];13(1):10.1517/14740338.2013.827660. Available from: https://pmc.ncbi.nlm.nih.gov/articles/PMC3864987/

10. Rochon PA, Gurwitz JH. The prescribing cascade revisited. The Lancet [Internet]. 2017 May 6 [cited 2025 Oct 14];389(10081):1778–80. Available from: https://www.thelancet.com/action/showFullText?pii=S0140673617311881

11. Hendrix MRS, Yasar M, Mohammad AK, Hugtenburg JG, Vanhommerig JW, Gündoğan-Yilmaz R, et al. Prescription Sequence Symmetry Analysis (PSSA) to assess prescribing cascades: a step-by-step guide. BMC Med Res Methodol [Internet]. 2024 Dec 1 [cited 2025 Oct 14];24(1):1–10. Available from: https://bmcmedresmethodol.biomedcentral.com/articles/10.1186/s12874-023-02108-y

12. Hallas J. Evidence of depression provoked by cardiovascular medication: A prescription sequence symmetry analysis. Epidemiology. 1996;7(5):478–84.

13. Janetzki JL, Sykes MJ, Ward MB, Pratt NL. Chronic Obstructive Pulmonary Disease Adverse Event Signals Associated with Potential Inhibitors of Glutathione Peroxidase 1: A Sequence Symmetry Analysis. Drug Saf [Internet]. 2024 Jan 1 [cited 2025 Oct 14];47(1):59–70. Available from: https://pubmed.ncbi.nlm.nih.gov/37995048/

14. Ndai AM, Smith K, Keshwani S, Choi J, Luvera M, Hunter J, et al. High-Throughput Screening for Prescribing Cascades Among Real-World Angiotensin-Converting Enzyme Inhibitor Initiators. Pharmacoepidemiol Drug Saf [Internet]. 2025 Mar 1 [cited 2025 Oct 14];34(3). Available from: https://pubmed.ncbi.nlm.nih.gov/40098294/

15. Doherty AS, Lund LC, Moriarty F, Boland F, Clyne B, Fahey T, et al. Prescribing Cascades Among Older Community-Dwelling Adults: Application of Prescription Sequence Symmetry Analysis to a National Database in Ireland. The Annals of Family Medicine [Internet]. 2025 Jun 10 [cited 2025 Oct 14];23(4):315–24. Available from: https://www.annfammed.org/content/early/2025/06/05/afm.240383

16. Lai ECC, Pratt N, Hsieh CY, Lin SJ, Pottegård A, Roughead EE, et al. Sequence symmetry analysis in pharmacovigilance and pharmacoepidemiologic studies. Eur J Epidemiol [Internet]. 2017 Jul 1 [cited 2025 Oct 14];32(7):567–82. Available from: https://pubmed.ncbi.nlm.nih.gov/28698923/

17. Sanchez-Santos MT, Axson EL, Dedman D, Delmestri A. Data Resource Profile Update: CPRD GOLD. Int J Epidemiol [Internet]. 2025 Aug 1 [cited 2025 Oct 14];54(4). Available from: https://pubmed.ncbi.nlm.nih.gov/40499193/

18. Voss EA, Makadia R, Matcho A, Ma Q, Knoll C, Schuemie M, et al. Feasibility and utility of applications of the common data model to multiple, disparate observational health databases. J Am Med Inform Assoc [Internet]. 2015 May 1 [cited 2025 Oct 14];22(3):553–64. Available from: https://pubmed.ncbi.nlm.nih.gov/25670757/

19. Chen X, Stanford T, Guo Y, Raventos B, Du M, Li X, et al. CohortSymmetry: An R package to perform sequence symmetry analysis using the OMOP common data model. medRxiv [Internet]. 2025 Nov 17 [cited 2025 Nov 18];2025.11.14.25340229. Available from: https://www.medrxiv.org/content/10.1101/2025.11.14.25340229v1

20. Tsiropoulos I, Andersen M, Hallas J. Adverse events with use of antiepileptic drugs: a prescription and event symmetry analysis. Pharmacoepidemiol Drug Saf [Internet]. 2009 [cited 2025 Oct 15];18(6):483–91. Available from: https://pubmed.ncbi.nlm.nih.gov/19326363/

21. Folch J, Busquets O, Ettcheto M, Sánchez-López E, Castro-Torres RD, Verdaguer E, et al. Memantine for the Treatment of Dementia: A Review on its Current and Future Applications. Journal of Alzheimer’s Disease [Internet]. 2018 [cited 2025 Oct 14];62(3):1223. Available from: https://pmc.ncbi.nlm.nih.gov/articles/PMC5870028/

22. Pedregal-Pascual P, Guarner-Argente C, Tan EH, Golozar A, Duarte-Salles T, Rosen AW, et al. Incidence and survival of colorectal cancer in the United Kingdom from 2000-2021: A population-based cohort study. American Journal of Gastroenterology [Internet]. 2025 [cited 2025 Nov 21]; Available from: https://journals.lww.com/ajg/fulltext/9900/incidence_and_survival_of_colorectal_cancer_in_the.1672.aspx

23. Corby G, Barclay NL, Tan EH, Burn E, Delmestri A, Duarte-Salles T, et al. Incidence, prevalence, and survival of lung cancer in the United Kingdom from 2000–2021: a population-based cohort study. Transl Lung Cancer Res [Internet]. 2024 Sep 30 [cited 2025 Nov 21];13(9):2187–201. Available from: https://tlcr.amegroups.org/article/view/90838/html

24. Colovic MB, Krstic DZ, Lazarevic-Pasti TD, Bondzic AM, Vasic VM. Acetylcholinesterase Inhibitors: Pharmacology and Toxicology. Curr Neuropharmacol [Internet]. 2013 Apr 25 [cited 2025 Oct 14];11(3):315. Available from: https://pmc.ncbi.nlm.nih.gov/articles/PMC3648782/

25. Ruangritchankul S, Chantharit P, Srisuma S, Gray LC. Adverse Drug Reactions of Acetylcholinesterase Inhibitors in Older People Living with Dementia: A Comprehensive Literature Review. Ther Clin Risk Manag [Internet]. 2021 [cited 2025 Oct 14];17:927. Available from: https://pmc.ncbi.nlm.nih.gov/articles/PMC8427072/

26. Li X, Zhang Y, Zhang L. The Roles of Acetylcholine and Muscarinic Receptors in Allergic Rhinitis: Mechanisms, Clinical Insights and Future Directions. Curr Allergy Asthma Rep [Internet]. 2025 Dec 1 [cited 2025 Oct 15];25(1). Available from: https://pubmed.ncbi.nlm.nih.gov/40932538/

27. Wong CW. Pharmacotherapy for Dementia: A Practical Approach to the Use of Cholinesterase Inhibitors and Memantine. Drugs Aging [Internet]. 2016 Jul 1 [cited 2025 Oct 15];33(7):451–60. Available from: https://link.springer.com/article/10.1007/s40266-016-0372-3

28. Vouri SM, Possinger MC, Usmani S, Solberg LM, Manini T. Evaluation of the Potential Acetylcholinesterase Inhibitor-Induced Rhinorrhea Prescribing Cascade. J Am Geriatr Soc [Internet]. 2020 Feb 1 [cited 2025 Oct 14];68(2):440–1. Available from: https://pubmed.ncbi.nlm.nih.gov/31625144/

29. Darreh-Shori T, Jelic V. Safety and tolerability of transdermal and oral rivastigmine in Alzheimer’s disease and Parkinson’s disease dementia. Expert Opin Drug Saf [Internet]. 2010 Jan 10 [cited 2025 Oct 15];9(1):167–76. Available from: https://pubmed.ncbi.nlm.nih.gov/20021294/

30. Farlow M, Veloso F, Moline M, Yardley J, Brand-Schieber E, Bibbiani F, et al. Safety and tolerability of donepezil 23 mg in moderate to severe Alzheimer’s disease. BMC Neurol [Internet]. 2011 May 25 [cited 2025 Oct 15];11. Available from: https://pubmed.ncbi.nlm.nih.gov/21612646/

31. Blume-Peytavi U, Kottner J, Sterry W, Hodin MW, Griffiths TW, Watson REB, et al. Age-Associated Skin Conditions and Diseases: Current Perspectives and Future Options. Gerontologist [Internet]. 2016 Apr 1 [cited 2025 Oct 15];56(Suppl_2):S230–42. Available from: 10.1093/geront/gnw003

32. Onyike CU. Psychiatric Aspects of Dementia. Continuum : Lifelong Learning in Neurology [Internet]. 2016 Apr 1 [cited 2025 Oct 15];22(2 Dementia):600. Available from: https://pmc.ncbi.nlm.nih.gov/articles/PMC5390928/

33. Bittner N, Funk CSM, Schmidt A, Bermpohl F, Brandl EJ, Algharably EEA, et al. Psychiatric Adverse Events of Acetylcholinesterase Inhibitors in Alzheimer’s Disease and Parkinson’s Dementia: Systematic Review and Meta-Analysis. Drugs Aging [Internet]. 2023 Nov 1 [cited 2025 Oct 15];40(11):953. Available from: https://pmc.ncbi.nlm.nih.gov/articles/PMC10600312/

34. Zhang NK, Zhang SK, Zhang LI, Tao HW, Zhang GW. The neural basis of neuropsychiatric symptoms in Alzheimer’s disease. Front Aging Neurosci. 2024 Dec 5;16:1487875.

35. Lai ECC, Wong MB, Iwata I, Zhang Y, Hsieh CY, Kao Yang YH, et al. Risk of pneumonia in new users of cholinesterase inhibitors for dementia. J Am Geriatr Soc [Internet]. 2015 May 1 [cited 2025 Oct 30];63(5):869–76. Available from: https://pubmed.ncbi.nlm.nih.gov/25912671/

36. Lampela P, Tolppanen AM, Tanskanen A, Tiihonen J, Hartikainen S, Taipale H. Anticholinergic Exposure and Risk of Pneumonia in Persons with Alzheimer’s Disease: A Nested Case-Control Study. Journal of Alzheimer’s Disease [Internet]. 2016 Nov 28 [cited 2025 Oct 30]; Available from: /doi/pdf/10.3233/JAD-160956?download=true

37. Masurkar PP, Chatterjee S, Sherer JT, Aparasu RR. Antimuscarinic Cascade Across Individual Cholinesterase Inhibitors in Older Adults with Dementia. Drugs Aging [Internet]. 2021 Jul 1 [cited 2025 Oct 15];38(7):593–602. Available from: https://pubmed.ncbi.nlm.nih.gov/34027602/

38. Rege S, Holmes HM, Aparasu RR. PMH20 Antimuscarinic Prescribing Cascade with Acetylcholinesterase Inhibitor Use Among a National Cohort of Older Adults with Alzheimer’s Disease. Value in Health [Internet]. 2021 Jun 1 [cited 2025 Oct 15];24:S131. Available from: https://www.valueinhealthjournal.com/action/showFullText?pii=S10983015210085 97

39. Alghamdi A, Bijlsma MJ, de Vos S, Schuiling-Veninga CCM, Bos JHJ, Hak E. Association between Incidence of Prescriptions for Alzheimer’s Disease and Beta-Adrenoceptor Antagonists: A Prescription Sequence Symmetry Analysis. Pharmaceuticals (Basel) [Internet]. 2023 Dec 1 [cited 2025 Oct 14];16(12). Available from: https://pubmed.ncbi.nlm.nih.gov/38139820/

40. O’Mahony D, Cherubini A, Guiteras AR, Denkinger M, Beuscart JB, Onder G, et al. STOPP/START criteria for potentially inappropriate prescribing in older people: version 3. Eur Geriatr Med [Internet]. 2023 Aug 1 [cited 2025 Oct 14];14(4):625–32. Available from: https://pubmed.ncbi.nlm.nih.gov/37256475/

41. Maher RL, Hanlon J, Hajjar ER. Clinical Consequences of Polypharmacy in Elderly. Expert Opin Drug Saf [Internet]. 2013 Jan [cited 2025 Oct 30];13(1):10.1517/14740338.2013.827660. Available from: https://pmc.ncbi.nlm.nih.gov/articles/PMC3864987/

42. Díaz-Gutiérrez MJ, Martínez-Cengotitabengoa M, Sáez de Adana E, Cano AI, Martínez-Cengotitabengoa MT, Besga A, et al. Relationship between the use of benzodiazepines and falls in older adults: A systematic review. Maturitas [Internet]. 2017 Jul 1 [cited 2025 Oct 30];101:17–22. Available from: https://pubmed.ncbi.nlm.nih.gov/28539164/

43. Johannes CB, Varas-Lorenzo C, McQuay LJ, Midkiff KD, Fife D. Risk of serious ventricular arrhythmia and sudden cardiac death in a cohort of users of domperidone: A nested case-control study. Pharmacoepidemiol Drug Saf [Internet]. 2010 Sep 1 [cited 2025 Oct 30];19(9):881–8. Available from: /doi/pdf/10.1002/pds.2016

44. Esumi S, Ushio S, Zamami Y. Polypharmacy in Older Adults with Alzheimer’s Disease. Medicina (B Aires) [Internet]. 2022 Oct 1 [cited 2025 Oct 30];58(10):1445. Available from: https://pmc.ncbi.nlm.nih.gov/articles/PMC9608980/

45. Sinnott SJ, Lin KJ, Wang S, Hallas J, Desai R, Schneeweiss S, et al. Evaluating signals generated in a large-scale sequence symmetry analyses: macrolides and heart failure; and NSAIDs and pneumonia. medRxiv [Internet]. 2020 Mar 22 [cited 2025 Oct 14];2020.03.19.20038596. Available from: https://www.medrxiv.org/content/10.1101/2020.03.19.20038596v1

46. Efjestad AS, Ihle-Hansen H, Hjellvik V, Engedal K, Blix HS. Drug Use before and after Initiating Treatment with Acetylcholinesterase Inhibitors. Dement Geriatr Cogn Dis Extra [Internet]. 2019 Jan 1 [cited 2025 Oct 14];9(1):196. Available from: https://pmc.ncbi.nlm.nih.gov/articles/PMC6528096/

47. Robinson M, Rowett D, Leverton A, Mabbott V. Changes in utilisation of anticholinergic drugs after initiation of cholinesterase inhibitors. Pharmacoepidemiol Drug Saf. 2009;18(8):659–64.

