## Supplement1 for "Prescription Cascades Associated with Acetylcholinesterase Inhibitors Use: A High-Throughput Sequence Symmetry Analysis"

### **Table S1: Anatomical Therapeutic Chemical (ATC) class attrition showing final ATC classes available for the study**

| Attrition Reason | n |
| --- | --- |
| Starting ATC classes | 909 |
| Removed classes with no record counts | 147 |
| Removed classes with less than 500 record counts | 95 |
| Removed A11 VITAMINS | 19 |
| Removed A12 MINERAL SUPPLEMENTS | 9 |
| Removed J07 VACCINES | 28 |
| Removed V VARIOUS ATC class | 84 |
| Removed class containing compounds | 17 |
| Removed N06DX other dementia drugs ATC class | 1 |
| **Included classes for analysis** | 509 |

### **Table S2: Drug ingredient attrition showing final drug ingredients available for the study**

| Attrition Reason | n |
| --- | --- |
| Starting Drug Ingredients | 31556 |
| Removed ingredients with no record counts | 29618 |
| Removed ingredients with <500 record counts | 725 |
| Removed dementia drug ingredients | 5 |
| **Included ingredient for analysis** | 1208 |

### **Table S3: Anatomical Therapeutic Chemical (ATC) classes showing negative signals with an 180-day initiation window**

| **ATC Class** | **ATC Class Name** | **Index, N (%)** | **Marker N, (%)** | **CSR (99% CI)** | **ASR (99% CI)** | **NSR** |
| --- | --- | --- | --- | --- | --- | --- |
| A05BA | liver therapy | 53 (34.4%) | 101 (65.6%) | 0.52 [0.34 - 0.81] | 0.57 [0.37 - 0.88] | 0.916 |
| A10BA | biguanides | 124 (38.3%) | 200 (61.7%) | 0.62 [0.46 - 0.83] | 0.65 [0.48 - 0.87] | 0.954 |
| A10BB | sulfonylureas | 58 (36.0%) | 103 (64.0%) | 0.56 [0.37 - 0.85] | 0.61 [0.40 - 0.93] | 0.921 |
| B01AF | direct factor xa inhibitors | 269 (43.2%) | 353 (56.8%) | 0.76 [0.62 - 0.94] | 0.63 [0.51 - 0.78] | 1.208 |
| B03AA | iron bivalent oral preparations | 778 (43.5%) | 1,012 (56.5%) | 0.77 [0.68 - 0.87] | 0.81 [0.71 - 0.91] | 0.955 |
| B03BA | vitamin b12 cyanocobalamin and analogues | 722 (36.0%) | 1,282 (64.0%) | 0.56 [0.50 - 0.63] | 0.62 [0.55 - 0.70] | 0.909 |
| B03BB | folic acid and derivatives | 803 (34.0%) | 1,557 (66.0%) | 0.52 [0.46 - 0.58] | 0.54 [0.49 - 0.61] | 0.95 |
| C03AA | thiazides plain | 224 (38.8%) | 353 (61.2%) | 0.63 [0.51 - 0.79] | 0.77 [0.61 - 0.96] | 0.827 |
| C08CA | dihydropyridine derivatives | 475 (42.8%) | 635 (57.2%) | 0.75 [0.64 - 0.87] | 0.80 [0.68 - 0.93] | 0.941 |
| C09AA | ace inhibitors plain | 373 (40.7%) | 543 (59.3%) | 0.69 [0.58 - 0.82] | 0.75 [0.63 - 0.89] | 0.915 |
| C10AA | hmg co a reductase inhibitors | 595 (36.6%) | 1,032 (63.4%) | 0.58 [0.50 - 0.66] | 0.62 [0.55 - 0.71] | 0.923 |
| H03AA | thyroid hormones | 155 (39.8%) | 234 (60.2%) | 0.66 [0.51 - 0.86] | 0.72 [0.55 - 0.93] | 0.926 |
| N02BA | salicylic acid and derivatives | 786 (40.1%) | 1,173 (59.9%) | 0.67 [0.59 - 0.75] | 0.76 [0.67 - 0.85] | 0.887 |
| N05AH | diazepines oxazepines thiazepines and oxepines | 790 (45.2%) | 956 (54.8%) | 0.83 [0.73 - 0.94] | 0.85 [0.75 - 0.96] | 0.974 |
| N06AB | selective serotonin reuptake inhibitors | 1,229 (40.3%) | 1,824 (59.7%) | 0.67 [0.61 - 0.74] | 0.71 [0.65 - 0.78] | 0.945 |
| N06AX | other antidepressants | 1,179 (45.6%) | 1,406 (54.4%) | 0.84 [0.76 - 0.93] | 0.86 [0.78 - 0.95] | 0.975 |

CSR: crude sequence ratio; ASR: adjusted sequence ratio; NSR: null sequence ratio; ATC: Anatomical Therapeutic Chemical

### **Figure S1: Sensitivity analysis comparing time windows of 180- versus 360-day initiation window for Anatomical Therapeutic Chemical (ATC) classes.**

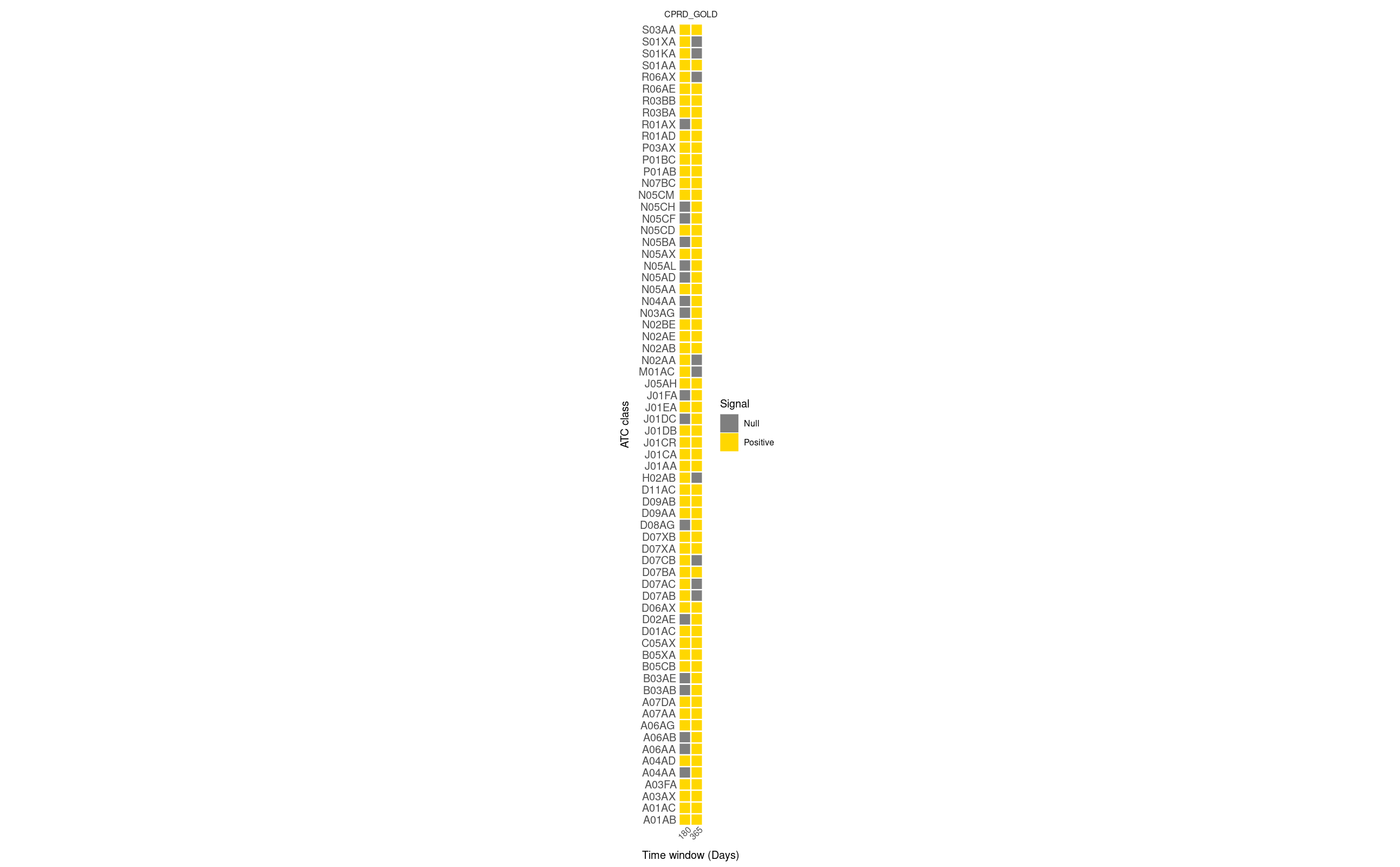

### **Table S4: Sensitivity analysis showing adjusted sequence ratios for time windows of 180 versus 360 days for Anatomical Therapeutic Chemical (ATC) classes (BOLD highlighting additional positive signals using a 365-day window).**

| Marker ATC class | ASR (99% CI) 180 days | ASR (99% CI) 365 days | signal classification (180 days) | signal classification (365 days) |
| --- | --- | --- | --- | --- |
| A01AB | 1.36 [1.04 - 1.78] | 1.31 [1.08 - 1.58] | Positive | Positive |
| A01AC | 2.38 [1.24 - 4.76] | 1.63 [1.01 - 2.64] | Positive | Positive |
| A03AX | 1.81 [1.22 - 2.70] | 1.35 [1.02 - 1.81] | Positive | Positive |
| A03FA | 1.58 [1.33 - 1.87] | 1.41 [1.24 - 1.60] | Positive | Positive |
| **A04AA** | **1.16 [0.57 - 2.37]** | **1.67 [1.01 - 2.81]** | **Null** | **Positive** |
| A04AD | 3.64 [2.43 - 5.59] | 4.34 [3.20 - 6.00] | Positive | Positive |
| **A06AA** | **1.11 [0.97 - 1.28]** | **1.15 [1.04 - 1.27]** | **Null** | **Positive** |
| **A06AB** | **1.30 [1.00 - 1.68]** | **1.25 [1.04 - 1.50]** | **Null** | **Positive** |
| A06AG | 1.33 [1.06 - 1.67] | 1.19 [1.01 - 1.40] | Positive | Positive |
| A07AA | 1.59 [1.27 - 2.00] | 1.62 [1.38 - 1.91] | Positive | Positive |
| A07DA | 1.50 [1.28 - 1.75] | 1.42 [1.27 - 1.59] | Positive | Positive |
| **B03AB** | **1.69 [0.72 - 4.17]** | **2.01 [1.04 - 4.03]** | **Null** | **Positive** |
| **B03AE** | **1.38 [0.98 - 1.94]** | **1.30 [1.02 - 1.67]** | **Null** | **Positive** |
| B05CB | 1.33 [1.11 - 1.59] | 1.39 [1.22 - 1.59] | Positive | Positive |
| B05XA | 1.43 [1.17 - 1.75] | 1.51 [1.30 - 1.75] | Positive | Positive |
| C05AX | 1.41 [1.20 - 1.64] | 1.48 [1.32 - 1.66] | Positive | Positive |
| D01AC | 1.24 [1.07 - 1.44] | 1.15 [1.03 - 1.28] | Positive | Positive |
| **D02AE** | **1.26 [0.91 - 1.74]** | **1.31 [1.04 - 1.64]** | **Null** | **Positive** |
| D06AX | 1.32 [1.11 - 1.57] | 1.15 [1.01 - 1.30] | Positive | Positive |
| D07AB | 1.20 [1.05 - 1.37] | 1.09 [0.99 - 1.20] | Positive | Null |
| D07AC | 1.16 [1.02 - 1.33] | 1.06 [0.96 - 1.16] | Positive | Null |
| D07BA | 1.25 [1.01 - 1.56] | 1.28 [1.09 - 1.51] | Positive | Positive |
| D07CB | 2.46 [1.05 - 6.16] | 1.33 [0.74 - 2.40] | Positive | Null |
| D07XA | 1.15 [1.02 - 1.29] | 1.12 [1.03 - 1.21] | Positive | Positive |
| D07XB | 1.48 [1.11 - 1.98] | 1.30 [1.06 - 1.59] | Positive | Positive |
| **D08AG** | **1.97 [0.99 - 4.09]** | **1.89 [1.13 - 3.22]** | **Null** | **Positive** |
| D09AA | 1.37 [1.15 - 1.64] | 1.20 [1.06 - 1.36] | Positive | Positive |
| D09AB | 1.44 [1.22 - 1.70] | 1.57 [1.39 - 1.77] | Positive | Positive |
| D11AC | 1.39 [1.22 - 1.58] | 1.35 [1.22 - 1.48] | Positive | Positive |
| H02AB | 1.19 [1.02 - 1.40] | 1.04 [0.93 - 1.16] | Positive | Null |
| J01AA | 1.17 [1.01 - 1.35] | 1.15 [1.04 - 1.28] | Positive | Positive |
| J01CA | 1.19 [1.07 - 1.32] | 1.13 [1.05 - 1.21] | Positive | Positive |
| J01CR | 1.21 [1.06 - 1.38] | 1.19 [1.09 - 1.31] | Positive | Positive |
| J01DB | 1.28 [1.11 - 1.47] | 1.18 [1.07 - 1.31] | Positive | Positive |
| **J01DC** | **1.39 [0.94 - 2.07]** | **1.54 [1.15 - 2.06]** | **Null** | **Positive** |
| J01EA | 1.10 [1.01 - 1.20] | 1.13 [1.06 - 1.20] | Positive | Positive |
| **J01FA** | **1.15 [1.00 - 1.32]** | **1.14 [1.03 - 1.26]** | **Null** | **Positive** |
| J05AH | 2.55 [1.05 - 6.81] | 2.84 [1.46 - 5.88] | Positive | Positive |
| M01AC | 1.64 [1.05 - 2.58] | 1.14 [0.83 - 1.56] | Positive | Null |
| N02AA | 1.15 [1.03 - 1.28] | 1.05 [0.97 - 1.14] | Positive | Null |
| N02AB | 2.67 [1.82 - 3.99] | 2.59 [1.97 - 3.44] | Positive | Positive |
| N02AE | 1.35 [1.08 - 1.70] | 1.38 [1.17 - 1.64] | Positive | Positive |
| N02BE | 1.14 [1.05 - 1.25] | 1.09 [1.02 - 1.16] | Positive | Positive |
| **N03AG** | **1.17 [0.81 - 1.69]** | **1.52 [1.14 - 2.01]** | **Null** | **Positive** |
| **N04AA** | **1.84 [0.99 - 3.49]** | **1.65 [1.07 - 2.57]** | **Null** | **Positive** |
| N05AA | 1.82 [1.42 - 2.34] | 2.53 [2.09 - 3.07] | Positive | Positive |
| **N05AD** | **1.14 [0.90 - 1.44]** | **1.54 [1.29 - 1.84]** | **Null** | **Positive** |
| **N05AL** | **1.04 [0.78 - 1.39]** | **1.31 [1.06 - 1.63]** | **Null** | **Positive** |
| N05AX | 1.17 [1.01 - 1.35] | 1.43 [1.27 - 1.61] | Positive | Positive |
| **N05BA** | **1.04 [0.93 - 1.16]** | **1.15 [1.06 - 1.25]** | **Null** | **Positive** |
| N05CD | 1.83 [1.55 - 2.16] | 2.23 [1.96 - 2.53] | Positive | Positive |
| **N05CF** | **0.97 [0.85 - 1.09]** | **1.13 [1.03 - 1.23]** | **Null** | **Positive** |
| **N05CH** | **1.21 [0.89 - 1.66]** | **1.37 [1.07 - 1.75]** | **Null** | **Positive** |
| N05CM | 2.77 [1.97 - 3.95] | 3.47 [2.67 - 4.57] | Positive | Positive |
| N07BC | 10.34 [5.31 - 22.68] | 15.99 [9.20 - 30.34] | Positive | Positive |
| P01AB | 1.44 [1.14 - 1.84] | 1.32 [1.11 - 1.57] | Positive | Positive |
| P01BC | 1.69 [1.34 - 2.14] | 1.44 [1.20 - 1.73] | Positive | Positive |
| P03AX | 2.40 [1.19 - 5.09] | 2.18 [1.28 - 3.79] | Positive | Positive |
| R01AD | 1.66 [1.33 - 2.08] | 1.39 [1.19 - 1.63] | Positive | Positive |
| **R01AX** | **1.18 [0.85 - 1.65]** | **1.30 [1.02 - 1.66]** | **Null** | **Positive** |
| R03BA | 1.54 [1.30 - 1.83] | 1.30 [1.15 - 1.47] | Positive | Positive |
| R03BB | 1.36 [1.02 - 1.82] | 1.33 [1.08 - 1.63] | Positive | Positive |
| R06AE | 1.66 [1.43 - 1.93] | 1.44 [1.29 - 1.60] | Positive | Positive |
| R06AX | 1.30 [1.03 - 1.65] | 1.06 [0.90 - 1.25] | Positive | Null |
| S01AA | 1.20 [1.05 - 1.38] | 1.21 [1.10 - 1.34] | Positive | Positive |
| S01KA | 1.27 [1.02 - 1.58] | 1.08 [0.92 - 1.27] | Positive | Null |
| S01XA | 1.22 [1.02 - 1.47] | 1.10 [0.96 - 1.25] | Positive | Null |
| S03AA | 1.23 [1.08 - 1.41] | 1.15 [1.04 - 1.27] | Positive | Positive |

ASR: adjusted sequence ratio; ATC: Anatomical Therapeutic Chemical

### **Table S5: Ingredients showing negative signals with an 180-day initiation window**

| **name** | **Index, N (%)** | **Marker, N (%)** | **CSR (99% CI)** | **ASR (99% CI)** | **NSR** |
| --- | --- | --- | --- | --- | --- |
| aspirin | 767 (39.7%) | 1,165 (60.3%) | 0.66 [0.58 - 0.74] | 0.74 [0.66 - 0.83] | 0.891 |
| vitamin b12 | 120 (26.8%) | 328 (73.2%) | 0.37 [0.28 - 0.48] | 0.34 [0.26 - 0.45] | 1.07 |
| bendroflumethiazide | 224 (38.8%) | 353 (61.2%) | 0.63 [0.51 - 0.79] | 0.77 [0.62 - 0.96] | 0.827 |
| clopidogrel | 413 (44.5%) | 515 (55.5%) | 0.80 [0.68 - 0.95] | 0.83 [0.70 - 0.98] | 0.969 |
| amlodipine | 449 (42.6%) | 605 (57.4%) | 0.74 [0.63 - 0.87] | 0.77 [0.66 - 0.91] | 0.958 |
| propranolol | 62 (38.0%) | 101 (62.0%) | 0.61 [0.40 - 0.93] | 0.62 [0.41 - 0.94] | 0.984 |
| perindopril | 68 (36.4%) | 119 (63.6%) | 0.57 [0.38 - 0.84] | 0.64 [0.43 - 0.94] | 0.891 |
| hydroxocobalamin | 453 (31.7%) | 974 (68.3%) | 0.47 [0.40 - 0.54] | 0.47 [0.40 - 0.54] | 0.996 |
| ferrous sulfate | 358 (42.6%) | 483 (57.4%) | 0.74 [0.62 - 0.89] | 0.82 [0.69 - 0.98] | 0.902 |
| levothyroxine | 154 (39.7%) | 234 (60.3%) | 0.66 [0.50 - 0.86] | 0.71 [0.54 - 0.93] | 0.926 |
| metformin | 124 (38.3%) | 200 (61.7%) | 0.62 [0.46 - 0.83] | 0.65 [0.48 - 0.87] | 0.954 |
| pioglitazone | 9 (25.7%) | 26 (74.3%) | 0.35 [0.12 - 0.89] | 0.38 [0.13 - 0.98] | 0.9 |
| simvastatin | 499 (37.9%) | 817 (62.1%) | 0.61 [0.53 - 0.71] | 0.69 [0.59 - 0.80] | 0.888 |
| atorvastatin | 433 (39.3%) | 668 (60.7%) | 0.65 [0.55 - 0.76] | 0.65 [0.55 - 0.76] | 1.001 |
| ferrous fumarate | 619 (44.9%) | 761 (55.1%) | 0.81 [0.71 - 0.93] | 0.81 [0.71 - 0.93] | 1.002 |
| gliclazide | 54 (36.0%) | 96 (64.0%) | 0.56 [0.36 - 0.87] | 0.61 [0.39 - 0.93] | 0.928 |
| cholecalciferol | 317 (42.2%) | 434 (57.8%) | 0.73 [0.60 - 0.88] | 0.65 [0.53 - 0.78] | 1.128 |
| folic acid | 744 (32.5%) | 1,542 (67.5%) | 0.48 [0.43 - 0.54] | 0.48 [0.43 - 0.54] | 0.997 |
| thiamine | 87 (32.3%) | 182 (67.7%) | 0.48 [0.34 - 0.67] | 0.49 [0.35 - 0.68] | 0.973 |
| rivaroxaban | 122 (44.9%) | 150 (55.1%) | 0.81 [0.59 - 1.11] | 0.69 [0.50 - 0.94] | 1.177 |
| almond oil | 62 (39.5%) | 95 (60.5%) | 0.65 [0.43 - 0.99] | 0.65 [0.43 - 0.99] | 1 |
| apixaban | 139 (42.8%) | 186 (57.2%) | 0.75 [0.56 - 1.00] | 0.61 [0.45 - 0.81] | 1.231 |
| edoxaban | 36 (40.0%) | 54 (60.0%) | 0.67 [0.38 - 1.15] | 0.53 [0.30 - 0.92] | 1.251 |
| duloxetine | 67 (38.3%) | 108 (61.7%) | 0.62 [0.41 - 0.92] | 0.60 [0.40 - 0.89] | 1.04 |
| mirtazapine | 909 (46.4%) | 1,051 (53.6%) | 0.86 [0.77 - 0.97] | 0.85 [0.76 - 0.96] | 1.012 |
| sertraline | 594 (41.0%) | 855 (59.0%) | 0.69 [0.61 - 0.80] | 0.66 [0.58 - 0.76] | 1.048 |
| venlafaxine | 101 (33.4%) | 201 (66.6%) | 0.50 [0.37 - 0.69] | 0.55 [0.40 - 0.74] | 0.921 |
| fluoxetine | 147 (34.2%) | 283 (65.8%) | 0.52 [0.40 - 0.67] | 0.56 [0.43 - 0.72] | 0.93 |
| quetiapine | 694 (45.2%) | 840 (54.8%) | 0.83 [0.72 - 0.94] | 0.83 [0.72 - 0.94] | 0.998 |
| citalopram | 795 (41.6%) | 1,114 (58.4%) | 0.71 [0.63 - 0.80] | 0.77 [0.68 - 0.87] | 0.928 |
| chlordiazepoxide | 7 (20.6%) | 27 (79.4%) | 0.26 [0.08 - 0.71] | 0.30 [0.09 - 0.82] | 0.866 |

CSR: crude sequence ratio; ASR: adjusted sequence ratio; NSR: null sequence ratio; ATC: Anatomical Therapeutic Chemical

### **Figure S2: Sensitivity analysis comparing time windows of 180 versus 360 days for drug ingredients.**

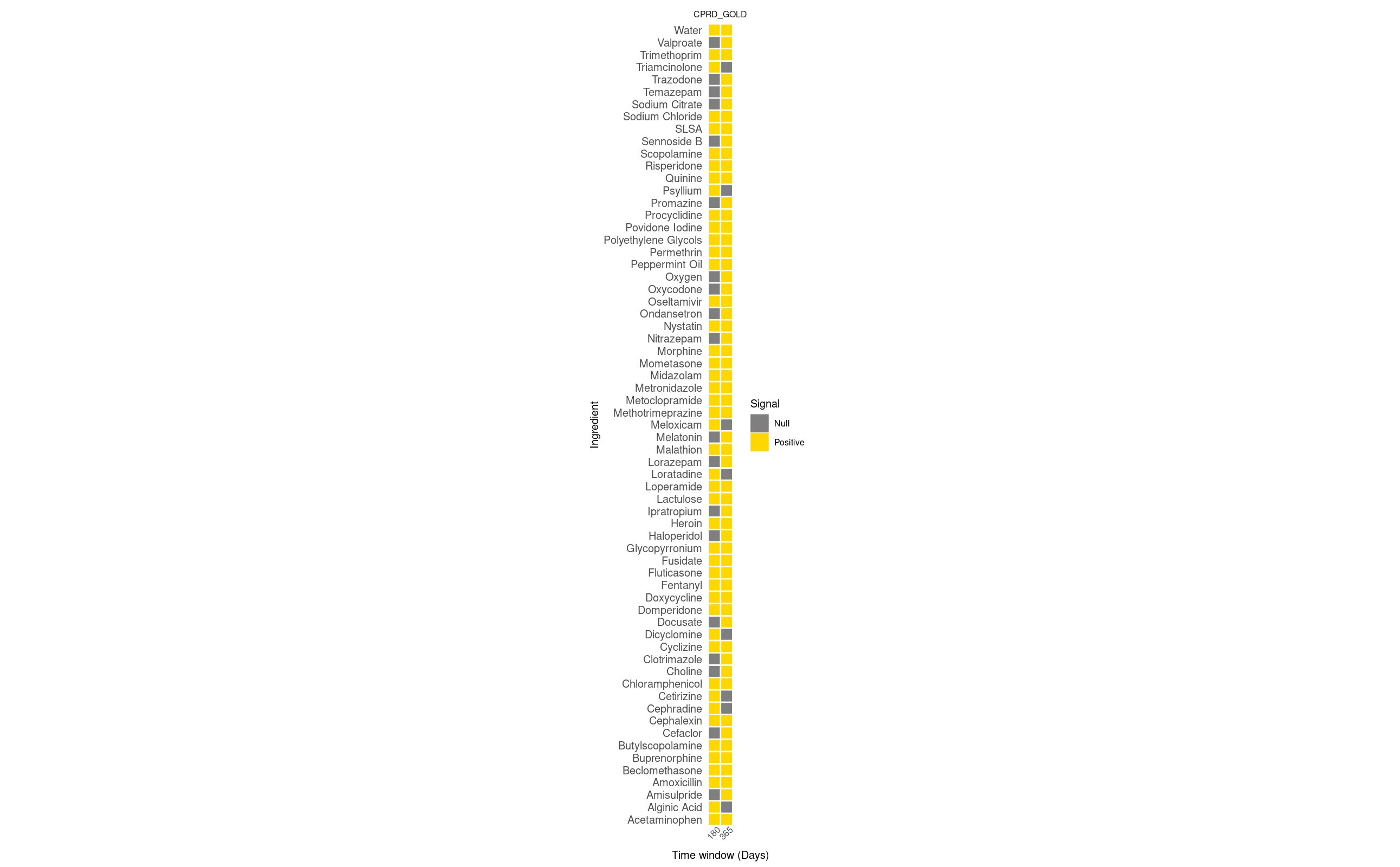

### **Table S6: Sensitivity analysis showing adjusted sequence ratios for time windows of 180 versus 365 days for ingredient (Bold highlighting additional positive signals using 365-days window).**

| Marker name | ASR (99% CI) 180 days | ASR (99% CI) 365 days | signal classification (180 days) | signal classification (365 days) |
| --- | --- | --- | --- | --- |
| **Ondansetron** | **1.15 [0.57 - 2.36]** | **1.66 [1.01 - 2.79]** | **Null** | **Positive** |
| **Clotrimazole** | **1.15 [0.97 - 1.36]** | **1.18 [1.04 - 1.33]** | **Null** | **Positive** |
| Loratadine | 1.41 [1.06 - 1.89] | 1.04 [0.85 - 1.28] | Positive | Null |
| Morphine | 2.54 [2.03 - 3.20] | 2.61 [2.22 - 3.09] | Positive | Positive |
| **Ipratropium** | **1.34 [0.89 - 2.03]** | **1.50 [1.13 - 2.01]** | **Null** | **Positive** |
| Beclomethasone | 1.48 [1.21 - 1.82] | 1.20 [1.04 - 1.40] | Positive | Positive |
| **Oxycodone** | **0.97 [0.63 - 1.49]** | **1.40 [1.03 - 1.92]** | **Null** | **Positive** |
| Acetaminophen | 1.17 [1.07 - 1.27] | 1.09 [1.02 - 1.16] | Positive | Positive |
| Buprenorphine | 1.35 [1.08 - 1.70] | 1.38 [1.17 - 1.64] | Positive | Positive |
| Cetirizine | 1.23 [1.02 - 1.49] | 1.12 [0.98 - 1.28] | Positive | Null |
| Fluticasone | 1.51 [1.14 - 1.99] | 1.34 [1.10 - 1.64] | Positive | Positive |
| Meloxicam | 1.66 [1.06 - 2.65] | 1.12 [0.81 - 1.55] | Positive | Null |
| Fentanyl | 2.75 [1.87 - 4.13] | 2.65 [2.01 - 3.53] | Positive | Positive |
| **Choline** | **1.30 [0.71 - 2.41]** | **1.65 [1.04 - 2.63]** | **Null** | **Positive** |
| **Melatonin** | **1.21 [0.89 - 1.67]** | **1.37 [1.07 - 1.75]** | **Null** | **Positive** |
| Trimethoprim | 1.10 [1.01 - 1.20] | 1.13 [1.06 - 1.20] | Positive | Positive |
| Metronidazole | 1.42 [1.15 - 1.76] | 1.27 [1.09 - 1.48] | Positive | Positive |
| Amoxicillin | 1.20 [1.08 - 1.33] | 1.14 [1.05 - 1.22] | Positive | Positive |
| Doxycycline | 1.19 [1.03 - 1.38] | 1.16 [1.04 - 1.29] | Positive | Positive |
| Povidone Iodine | 2.61 [1.23 - 5.86] | 2.47 [1.43 - 4.40] | Positive | Positive |
| Quinine | 1.68 [1.33 - 2.12] | 1.43 [1.19 - 1.72] | Positive | Positive |
| **Cefaclor** | **1.35 [0.88 - 2.07]** | **1.46 [1.07 - 2.00]** | **Null** | **Positive** |
| Cephalexin | 1.28 [1.10 - 1.48] | 1.19 [1.06 - 1.32] | Positive | Positive |
| Cephradine | 1.56 [1.03 - 2.39] | 1.21 [0.88 - 1.68] | Positive | Null |
| Oseltamivir | 2.55 [1.05 - 6.81] | 2.78 [1.42 - 5.77] | Positive | Positive |
| Methotrimeprazine | 18.18 [8.25 - 48.64] | 25.73 [13.56 - 55.48] | Positive | Positive |
| Water | 8.47 [5.07 - 15.11] | 10.72 [7.34 - 16.25] | Positive | Positive |
| Fusidate | 1.29 [1.10 - 1.51] | 1.18 [1.05 - 1.32] | Positive | Positive |
| SLSA | 2.56 [1.40 - 4.83] | 1.73 [1.15 - 2.62] | Positive | Positive |
| **Nitrazepam** | **1.82 [1.00 - 3.38]** | **2.02 [1.27 - 3.28]** | **Null** | **Positive** |
| Heroin | 13.27 [6.17 - 33.86] | 21.44 [11.25 - 46.37] | Positive | Positive |
| **Oxygen** | **2.53 [0.84 - 8.81]** | **2.86 [1.25 - 7.21]** | **Null** | **Positive** |
| Alginic Acid | 1.58 [1.01 - 2.49] | 1.19 [0.86 - 1.63] | Positive | Null |
| Domperidone | 1.54 [1.24 - 1.91] | 1.31 [1.12 - 1.54] | Positive | Positive |
| **Promazine** | **1.01 [0.69 - 1.48]** | **1.39 [1.03 - 1.87]** | **Null** | **Positive** |
| **Amisulpride** | **1.01 [0.75 - 1.37]** | **1.30 [1.04 - 1.64]** | **Null** | **Positive** |
| Peppermint Oil | 2.00 [1.30 - 3.14] | 1.44 [1.05 - 1.99] | Positive | Positive |
| **Sennoside B** | **1.07 [0.94 - 1.21]** | **1.12 [1.02 - 1.22]** | **Null** | **Positive** |
| Butylscopolamine | 2.30 [1.76 - 3.04] | 2.19 [1.82 - 2.65] | Positive | Positive |
| Glycopyrronium | 4.39 [2.37 - 8.77] | 5.29 [3.35 - 8.74] | Positive | Positive |
| **Trazodone** | **1.15 [0.93 - 1.41]** | **1.44 [1.23 - 1.69]** | **Null** | **Positive** |
| Midazolam | 11.20 [6.89 - 19.39] | 16.54 [11.11 - 25.72] | Positive | Positive |
| Risperidone | 1.21 [1.03 - 1.41] | 1.48 [1.31 - 1.68] | Positive | Positive |
| Valproate | 1.17 [0.81 - 1.69] | 1.51 [1.14 - 2.01] | Null | Positive |
| Procyclidine | 2.00 [1.03 - 4.06] | 1.88 [1.18 - 3.04] | Positive | Positive |
| **Haloperidol** | **1.13 [0.89 - 1.42]** | **1.53 [1.28 - 1.83]** | **Null** | **Positive** |
| **Lorazepam** | **1.11 [0.97 - 1.29]** | **1.58 [1.41 - 1.77]** | **Null** | **Positive** |
| **Temazepam** | **1.15 [0.94 - 1.41]** | **1.29 [1.11 - 1.51]** | **Null** | **Positive** |
| Triamcinolone | 1.61 [1.06 - 2.47] | 1.13 [0.83 - 1.53] | Positive | Null |
| Mometasone | 1.30 [1.05 - 1.62] | 1.19 [1.01 - 1.39] | Positive | Positive |
| Metoclopramide | 1.57 [1.25 - 1.99] | 1.44 [1.21 - 1.72] | Positive | Positive |
| Cyclizine | 2.10 [1.72 - 2.59] | 1.88 [1.61 - 2.19] | Positive | Positive |
| Nystatin | 1.58 [1.26 - 1.99] | 1.62 [1.38 - 1.92] | Positive | Positive |
| Permethrin | 2.16 [1.38 - 3.45] | 2.14 [1.56 - 2.97] | Positive | Positive |
| Dicyclomine | 3.55 [1.24 - 12.10] | 1.60 [0.79 - 3.29] | Positive | Null |
| **Docusate** | **1.18 [0.93 - 1.50]** | **1.21 [1.02 - 1.45]** | **Null** | **Positive** |
| Polyethylene Glycols | 2.09 [1.18 - 3.82] | 1.87 [1.22 - 2.89] | Positive | Positive |
| Psyllium | 1.46 [1.05 - 2.05] | 1.03 [0.82 - 1.30] | Positive | Null |
| Scopolamine | 3.63 [2.43 - 5.59] | 4.39 [3.23 - 6.08] | Positive | Positive |
| Sodium Chloride | 1.27 [1.08 - 1.51] | 1.34 [1.18 - 1.52] | Positive | Positive |
| **Sodium Citrate** | **1.20 [0.87 - 1.68]** | **1.34 [1.05 - 1.71]** | **Null** | **Positive** |
| Lactulose | 1.17 [1.04 - 1.33] | 1.11 [1.01 - 1.21] | Positive | Positive |
| Chloramphenicol | 1.27 [1.10 - 1.47] | 1.23 [1.11 - 1.37] | Positive | Positive |
| Loperamide | 1.52 [1.30 - 1.77] | 1.44 [1.29 - 1.62] | Positive | Positive |
| Malathion | 2.40 [1.12 - 5.46] | 2.36 [1.33 - 4.32] | Positive | Positive |

### **Table S7: Review of positive signals for Anatomical Therapeutic Chemical (ATC) classes using an 180-day window**

See supplement excel file

### **Table S8: Review of positive signals for ingredient using a180-day window**

See supplement excel file

### **Table S9: Review of additional positive signals for Anatomical Therapeutic Chemical (ATC) classes using a 365-day window**

See supplement excel file

### **Table S10: Review of additional positive signals for ingredient level using a 365-day window**

See supplement excel file
